# NEO-EXCEL: Neoadjuvant trial of pre-operative exemestane or letrozole, with or without celecoxib, in the treatment of oestrogen receptor-positive postmenopausal early breast cancer: A phase III, randomised, double-blind, placebo-controlled trial

**DOI:** 10.64898/2026.07.13.26356308

**Authors:** Adele Francis, Amit Patel, Sarah J. Pirrie, Catharine Prest, Cassey L. Brookes, John M. S. Bartlett, Robert C. Stein, Janet A. Dunn, Peter Canney, Christopher J. Poole, Ashraf R. Patel, Margaret Grant, Katy Herring, Elizabeth Southgate, Claire Gaunt, Sarah J. Bowden, Daniel W. Rea

## Abstract

**Background:** The NEO-EXCEL trial hypothesised that aromatase inhibitor (AI)-activity as neoadjuvant endocrine therapy for early-stage breast cancer in postmenopausal women may be enhanced in combination with cyclooxygenase-2 (COX-2) inhibition.

**Methods:** NEO-EXCEL was a phase III, placebo-controlled, randomised trial in postmenopausal women with oestrogen receptor (ER)-positive resectable breast cancer with tumours ≥2cm. Women were randomised (1:1:1:1): exemestane (25mg od) plus celecoxib (400mg bid), exemestane (25mg od) plus placebo (bid), letrozole (2.5mg od) plus celecoxib (400mg bid), or letrozole (2.5mg od) plus placebo (bid). Primary endpoint was clinical response (complete/partial) measured by callipers at 16 weeks; a standard assessment method at the time of trial inception. Sixteen-week ultrasound-determined response was the main secondary outcome to verify the calliper-based primary. Analysis was intention-to-treat.

**Results:** Due to slow accrual the trial design was redesigned from a definitive 2×2, 1000 patient trial to one randomising 269 patients between 20-Nov-2007 and 29-Apr-2014; 34.9% were human epithelial growth factor receptor 2-positive. AI+celecoxib produced a significantly greater objective clinical response than AI+placebo (72.9% vs 55.6%, *P*=0.003), which remained after adjustment for AI type and stratification factors (odds ratio = 2.3; 95% CI 1.3-3.8, *P*=0.003). Ultrasound-determined response was however not significantly enhanced (48.7% [AI+celecoxib] vs 41.2% [AI+placebo], *P*=0.34). Progression free survival and overall survival remained similar (median follow-up = 5.1 years [range 0.1-7.1]).

**Conclusions:** NEO-EXCEL is the first completed, phase III double-blind, placebo-controlled trial testing the addition of celecoxib to AI as neoadjuvant endocrine therapy in early breast cancer. Clinical response showed significant improvement but there was no significant ultrasound-determined response improvement nor any surgical or long-term outcome evidence of AI+COX-2 inhibition improving treatment outcomes for ER+ early resectable postmenopausal breast cancers. Use of short-term celecoxib at 400mg bd for 16 weeks was safe with no excess cardiotoxicity observed.

**Trial registration:** EudraCT Number: 2006-000436-27; 16-Feb-2007

ISRCTN number: 09768535; 11-Jul-2007

## Introduction

Whilst many women will remain disease free following diagnosis with an early hormone receptor positive (HR+) breast cancer, approximately a third will relapse. Response rates in postmenopausal women to contemporary neoadjuvant chemotherapy regimens show similar activity to endocrine therapy to those with HR+ cancers in terms of clinical or pathological complete response [1, 2]. In postmenopausal women aromatase inhibition appears more effective than tamoxifen in reducing tumour size and in down-staging to permit breast conservation therapy [3].

To date, no meaningful differences in clinical activity between aromatase inhibitors (AIs) have emerged, with similar activity demonstrated in head-to-head comparisons in the neoadjuvant, adjuvant and advanced disease settings [4–7]. Neoadjuvant aromatase inhibition provides a safe, low-toxicity option for surgically down-staging postmenopausal women with oestrogen-receptor positive (ER+) breast cancer. Increasing the effectiveness of such therapy with additional agents would represent a significant advance [8].

Cyclooxygenase-2 (COX-2) is overexpressed in breast cancer and has been implicated in breast tumorigenesis, tumour proliferation, and invasion [9]. A review of COX-2 inhibitors in breast cancer has provided strong evidence from molecular, animal, and cell line studies that supports the ability of COX-2 inhibitors to prevent the development of breast tumours [10–12].

COX-2 and aromatase expression have been found to correlate in breast cancer tissue. COX-2 product prostaglandin E2 and cytokines such as interleukin-6 can upregulate aromatase expression [13]. Dual inhibition of aromatase and COX-2 may, therefore, synergise in controlling endocrine responsive breast cancer [14], with additional COX-2 mediated anticancer activity reported [15]. Although there are major cardiac safety concerns regarding long-term use of some COX-2 inhibitors, particularly in patients with significant cardiac risk factors, short-term use of celecoxib at the dose selected for this study (400mg twice daily [bid]) has an established acceptable cardiac safety profile in patients with low cardiovascular risk [16].

NEO-EXCEL was a phase III trial hypothesising that AI activity, as primary neoadjuvant therapy for early-stage ER+ postmenopausal breast cancers, may be enhanced by the addition of the COX-2 inhibitor celecoxib.

Response in the neoadjuvant setting is reported by varying methods between trials. At the time of NEO-EXCEL’s development there was accumulating evidence linking various measures of primary tumour response to eventual disease-free survival (DFS) with multiple examples of neoadjuvant endpoints predictive of long-term outcomes [8, 17]. To ensure comparison with other large endocrine trials performed in the neo-adjuvant setting [3, 8], clinical response in terms of reduction in palpable tumour was selected as the appropriate measure of primary tumour response in the NEO-EXCEL trial. Cleator et al [18], in their analysis of neoadjuvant chemoendocrine therapy showed good clinical response to be significantly associated with superior progression-free survival (PFS) and overall survival (OS). In addition, at a similar time, the PROACT trial demonstrated clinically beneficial tumour downstaging and reductions in tumour volume by ultrasound and calliper at baseline and at three months [19]. With two later trials also combining calliper measurements with imaging modalities [20, 21]. Although NEO-EXCEL included the key secondary assessment of using ultrasound measurements to provide supporting validation, we note that the primary outcome measure once standard within neoadjuvant endocrine studies is now deemed a non-standard assessment method.

Publication of these results were delayed so as to include the five-year outcome data, with analysis also delayed by the COVID-19 pandemic.

## Methods

### Trial design

NEO-EXCEL was a phase III, multicentre, randomised, placebo-controlled clinical trial conducted at 22 UK hospitals.

NEO-EXCEL was approved by the West Midlands Research Ethics Committee (ref: 06/MRE07/31, dated 24-Jul-2006) and conducted in accordance with the principles of the Declaration of Helsinki and Good Clinical Practice. Patients gave written informed consent. The current version of the protocol (v9.0, 09-Nov-2018) can be found here: https://www.birmingham.ac.uk/research/crctu/trials/neo-excel/index.aspx

The original design of NEO-EXCEL was ambitious; it was the largest neoadjuvant breast cancer trial in the UK at the time, aiming to open in 70 UK hospitals and estimating that 1000 patients were needed for a four-group comparison: exemestane plus celecoxib; exemestane plus placebo; letrozole plus celecoxib; and letrozole plus placebo. This allowed for the detection of differences not only between celecoxib and placebo but also aimed to determine superiority of exemestane compared to letrozole in combination with the COX-2 inhibitor. As per the protocol, an independent data monitoring committee (DMC) reviewed interim data of the first 50 patients randomised on 11-Dec-2009 (two years after trial opening); concerns were raised regarding slow trial accrual (see Discussion for more details). A protocol amendment was proposed and a new recruitment target of 256 was approved in protocol v4.0 (dated, 01-Feb-2010). The trial’s objective was thus simplified to only determine whether primary neo-adjuvant endocrine therapy for early breast stage ER+ cancer in postmenopausal women may be enhanced by the addition of the selective COX-2 inhibitor, celecoxib.

### Patients

Eligible patients were treatment naïve, postmenopausal women with biopsy proven, ER+ (Allred-Quick Score ≥3), invasive breast cancer tumours of ≥2cm diameter; Eastern Cooperative Oncology Group status 0-2; human epithelial growth factor receptor (HER)2 negative or positive (as per standard local methods at the time, predominantly immunohistochemistry with fluorescent in situ hybridisation). Postmenopausal was defined as bilateral surgical oophorectomy, amenorrhea at any age for ≥5 years (any cause; levonorgestrel devices were not formally excluded but unlikely) or natural amenorrhea for ≥1 year in women above 50 years. Patients with T4 tumours and any nodal involvement were permitted.

Exclusion criteria included ongoing requirements for non-steroidal anti-inflammatory drugs or COX-2 inhibitor therapy (aspirin 75mg od permissible); regular COX-2 inhibitor use in the two years prior to randomisation; inflammatory bowel disease; ongoing requirements for contraindicated concomitant medications including systemic oestrogen therapy, fluconazole and ketoconazole, angiotensin-converting enzyme inhibitors, and lithium. Patients discontinued hormone replacement therapies prior to trial entry.

### Randomisation and masking

Randomisation was 1:1:1:1 to either exemestane or letrozole with concurrent celecoxib, or exemestane or letrozole with placebo. Treatment allocation was performed by central randomisation at the CRCTU, University of Birmingham. Randomisation was stratified by tumour size (2-5cm, >5cm), grade (I, II, III), ER Quick Score (3-4, 5-6, 7-8), HER2 status (negative, positive, not determined), and low-dose aspirin use (no, yes). A standard minimisation procedure was used to ensure balance between the arms and stratification variables.

### Interventions and procedures

Patients received a 16-week neoadjuvant regime of either exemestane (25mg once daily [od]) or letrozole (2.5mg od) with either concurrent celecoxib (400mg bid orally) or placebo (bid) as outpatients. Dose modifications of trial treatment were not permitted.

Pre-treatment evaluation included: uni-dimensional tumour measurement by callipers of the longest dimension; tumour assessment by ultrasound; full blood count; urea, creatinine, and electrolytes; liver function tests, blood pressure measurement; and NYHA classification.

Uni-dimensional tumour measurements were performed 4-weekly while patients were on treatment. On treatment completion, tumour measurements were performed with a tumour ultrasound and repeated blood tests. Response was assessed by Response Evaluation Criteria In Solid Tumours (RECIST) version 1.0 [22].

Postoperative management depended on operative pathology and was determined following local protocol. It was expected that patients should receive a total of five years endocrine therapy; continuation of AI therapy was recommended.

Patients were followed up yearly for five years post-surgery and then as per local policies.

### Outcomes

The primary efficacy endpoint determined at 16 weeks consisted of either a partial response (PR) or complete response (CR) as measured by callipers; a standard assessment method at the time of the trial’s inception. CR was defined as complete disappearance of the radiological lesion (no palpable lesion detected), with PR a decrease in the longest uni-dimensional measurement by callipers of at least 30%. All other patients are classified as non-responders.

Objective ultrasound-determined response determined in patients achieving CR or PR at 16 weeks was the main secondary endpoint as assessed by RECIST v1.0 [22]. Patients recorded as not achieving either CR or PR at 16 weeks were defined as non-responders.

Other secondary endpoints were: type of surgery (breast conserving surgery or mastectomy); axillary lymph node involvement at surgery, yes or no; pathological response, defined as when all detectable invasive tumours had disappeared; local recurrence-free survival, defined as time from trial entry date to date when local recurrence was first observed; PFS, defined as the time from the trial entry date to date when progression was first observed; and OS, defined as the time from trial entry date to date of deaths from any cause. Patients were censored at date last seen in all time-to-event analyses.

The incidence of adverse events (AEs) graded 1-5 according to National Cancer Institute (NCI) – Common Terminology Criteria for Adverse Events (CTCAE) v3.0 are also reported, where Grade 1-to-3 correspond to mild, moderate, and severe AEs, respectively.

### Statistical analysis

The adjusted sample size of 256 analysable patients was powered for the main comparison of AI+celecoxib versus AI+placebo only. The trial was designed to provide 80% power, randomising 256 analysable patients to allow detection of absolute differences in response rates of >15% between AI+celecoxib and AI+placebo at a 10% (2-sided) significance level using a chi-squared test. Additional patients were recruited to allow for a small number who had unmeasurable disease at baseline or missing outcome data, resulting in recruitment of 269 patients. Analysis was performed on an intention-to-treat basis. There were no formal stopping rules.

Summary statistics (median, interquartile range [IQR]) were calculated for tumour size. The proportion of responders with 95% confidence intervals (CI) was calculated for response outcome measures. Response rates between AI+celecoxib and AI+placebo are compared after adjusting for type of AI and stratification variables in a logistic regression analysis. An unplanned logistic regression analysis was carried out to assess the treatment interaction of the primary outcome.

All secondary binary outcome measures were compared between primary treatment groups (AI+celecoxib versus AI+placebo) using chi-squared tests.

The database was locked on 16-Mar-2021. Analyses were performed using R version 4.0.3 or STATA v18.0.

As well as the interim 50-patient assessment described in the Trial Design section, the independent DMC reviewed the trial annually to ensure patient safety.

## Results

Between 20-Nov-2007 and 29-Apr-2014, 269 women, were randomly assigned to one of four treatment groups: 67 (24.9%) Placebo/Exemestane; 66 (24.5%) Placebo/Letrozole; 67 (24.9%) Celecoxib/Exemestane; or 69 (25.7%) Celecoxib/Letrozole (Figure 1). Three patients were excluded from the primary analysis of tumour response; two due to missing measurements at baseline and the third due to an unmeasurable tumour (<2cm) at baseline as defined by RECIST v1.0 [22], All of these received celecoxib. Three further patients did not start treatment, one each in the Placebo/Exemestane, Placebo/Letrozole, and Celecoxib/Exemestane arms. Baseline patient characteristics were comparable between the treatment groups, with an unexpectedly high proportion of patients with ER+HER2+ tumours randomised (34.9%; Table 1). Median age was 67 years (range 48-90).

**Figure 1.**
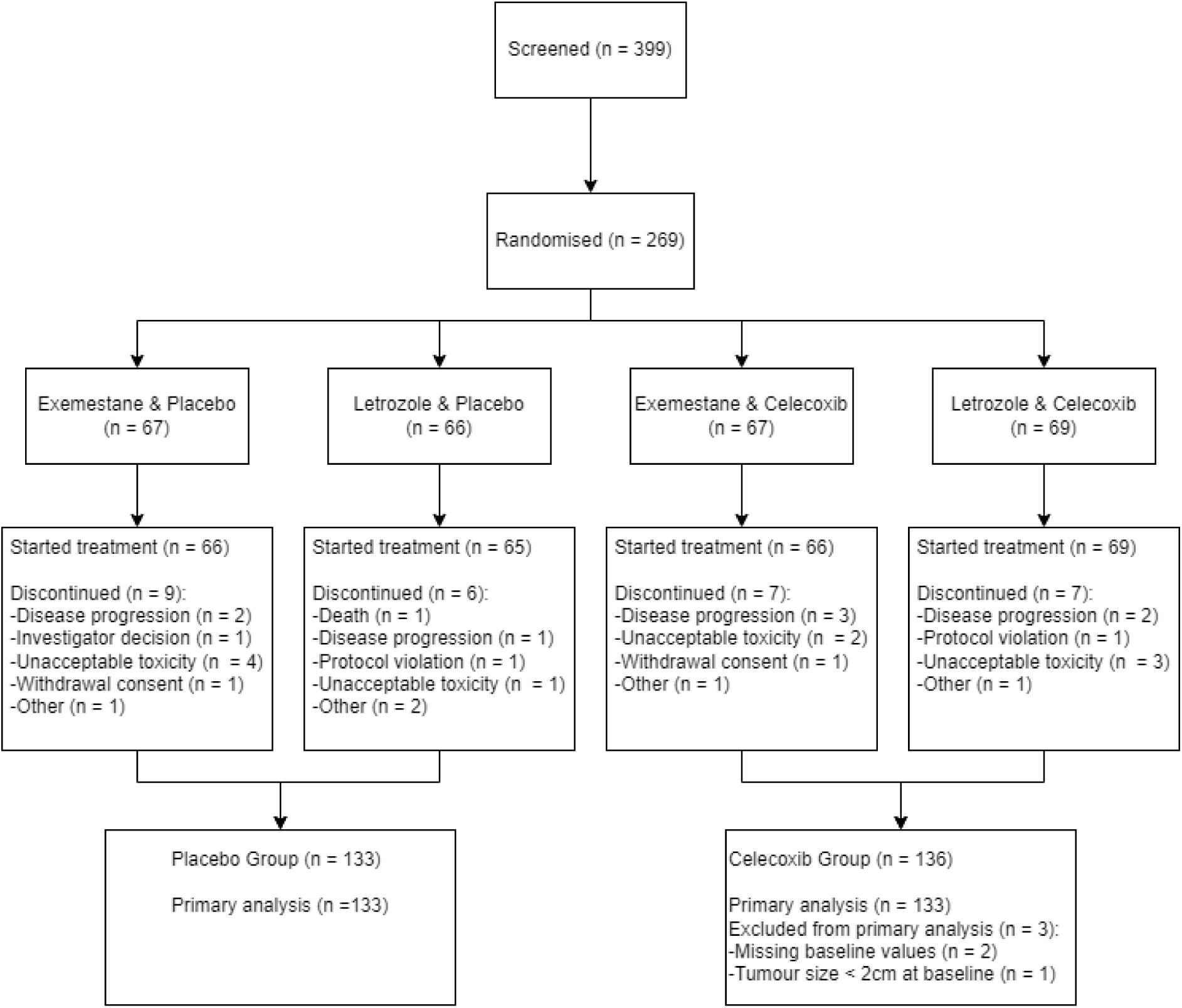
NEO-EXCEL trial profile. The number of patients included within the NEO-EXCEL trial are shown as per CONSORT.

**Table 1.** Baseline patient characteristics.

| Characteristic | Placebo [N (%)] |  | Celecoxib [N (%)] |  | Overall<br>N = 269 |
| --- | --- | --- | --- | --- | --- |
|  | Exemestane<br>N = 67 | Letrozole<br>N = 66 | Exemestane<br>N = 67 | Letrozole<br>N = 69 |  |
| **Age (years)** |  |  |  |  |  |
| Median | 67 | 64 | 66 | 71 | 67 |
| Range | 51, 86 | 48, 88 | 49, 90 | 50, 88 | 48, 90 |
| **ER Quick Score** |  |  |  |  |  |
| 3-4 | 1 (1.5) | 0 (0) | 2 (3) | 0 (0) | 3 (1.1) |
| 5-6 | 2 (3) | 1 (1.5) | 1 (1.5) | 3 (4.3) | 7 (2.6) |
| 7-8 | 64 (95.5) | 65 (98.5) | 64 (95.5) | 66 (95.7) | 259 (96.3) |
| **HER2** |  |  |  |  |  |
| Negative | 39 (58.2) | 40 (60.6) | 40 (59.7) | 41 (59.4) | 160 (59.5) |
| Positive | 24 (35.8) | 23 (34.8) | 23 (34.3) | 24 (34.8) | 94 (34.9) |
| Not determined | 4 (6) | 3 (4.5) | 4 (6) | 4 (5.8) | 15 (5.6) |
| **Tumour Grade** |  |  |  |  |  |
| Well-differentiated | 10 (14.9) | 8 (12.1) | 9 (13.4) | 9 (13) | 36 (13.4) |
| Moderately differentiated | 46 (68.7) | 46 (69.7) | 46 (68.7) | 47 (68.1) | 185 (68.8) |
| Poorly differentiated | 11 (16.4) | 12 (18.2) | 12 (17.9) | 13 (18.8) | 48 (17.8) |
| **Tumour Size** |  |  |  |  |  |
| >2-5cm | 55 (82.1) | 56 (84.8) | 58 (86.6) | 53 (76.8) | 222 (82.5) |
| >5cm | 12 (17.9) | 10 (15.2) | 9 (13.4) | 16 (23.2) | 47 (17.5) |
| **Aspirin Use** |  |  |  |  |  |
| No | 58 (86.6) | 59 (89.4) | 59 (88.1) | 60 (87) | 236 (87.7) |
| Yes | 9 (13.4) | 7 (10.6) | 8 (11.9) | 9 (13) | 33 (12.3) |

ER, oestrogen receptor; HER2, human epithelial growth factor receptor 2.

Twenty-nine patients discontinued trial treatment early. The most common reasons were unacceptable side effects (10 patients) and disease progression (8 patients). Treatment adherence remained high during the trial; 299 (85.4%) recorded taking all prescribed medication during four of the five two-weekly on-treatment visits (52 (77.6%) Placebo/Exemestane; 58 (89.2%) Placebo/Letrozole; 57 (85.1%) Celecoxib/Exemestane; and 62 (89.9%) Celecoxib/Letrozole).

Clinical response rates were calculated for 266 women, 22 of whom were categorised as ‘non-responders’ as per the intention-to-treat analysis because the 16-week tumour measurements were not available (10 receiving AI+celecoxib and 12 receiving AI+placebo). The AI+placebo treatment arm had a response rate of 55.6% (95% CI 46.8-64.3) and the AI+celecoxib arm 72.9% (95% CI 64.6-80.3). This increase was statistically significant (χ^2^ = 8.662 *P*=0.003). Response rates split by the four-arm treatment comparison are included in Table 2. Exploratory analysis using a univariate logistic regression model confirmed an increased odds ratio (OR) of 2.2 (95% CI 1.3-3.6, *P*=0.003) with celecoxib treatment. Additional adjustments for stratification factors and adjusting for the effect of AI treatment in a multivariate logistic regression also confirmed the potential benefit of celecoxib treatment (Appendix A).

**Table 2.** Clinical response rates by four-arm treatment comparison.

|  | Exemestane [*N* (%)] |  |  | Letrozole [*N* (%)] |  |  |
| --- | --- | --- | --- | --- | --- | --- |
|  | Placebo | Celecoxib | Total | Placebo | Celecoxib | Total |
| No response | 34 (50.8) | 15 (22.4) | 49 (36.6) | 25 (37.9) | 21 (31.8) | 46 (34.9) |
| Response | 33 (49.3) | 52 (77.6) | 85 (63.4) | 41 (62.1) | 45 (68.2) | 86 (65.2) |
| Total | 67 | 67 | 134 | 66 | 66 | 132 |

The median tumour size measured by callipers at baseline was 3.5cm in both AI+celecoxib and AI+placebo treatment groups (*N*=266, IQR 30‒45). After 16 weeks treatment, calliper measurements were available for 244 patients and decreased to 1.7cm (*N*=123, IQR 0.8-2.6) in the AI+celecoxib group and 2.0cm (*N*=121, IQR 1.3‒3.1) in the AI+placebo group.

The secondary treatment outcomes are presented in Table 3. Median tumour size as measured by ultrasound at baseline was smaller overall compared with calliper measurements (3.0cm vs 3.5cm). At completion of treatment, 213 ultrasounds were evaluable and tumour size decreased to 1.9cm (*N*=102, IQR 1.4-2.6) in the AI+celecoxib group, and 2.1cm (*N*=91, IQR 1.5‒3.0) in the AI+placebo group. Patients treated with AI+celecoxib had a statistically non-significant but numerically increased ultrasound-determined response rate compared to placebo, but the rate of breast conserving surgery and percentage of women with axillary lymph node involvement at surgery and pathological response is comparable between the AI+celecoxib and AI+placebo groups (Table 4).

**Table 3.** Secondary treatment outcomes.

|  | Placebo [*N* (%)] |  | Celecoxib [*N* (%)] |  |  |
| --- | --- | --- | --- | --- | --- |
|  | Exemestane | Letrozole | Exemestane | Letrozole | Overall |
| Objective Ultrasound-determined Response |  |  |  |  |  |
| No response | 32 (60.4) | 28 (57.1) | 25 (48.1) | 32 (54.2) | 117 (54.9) |
| Response | 21 (39.6) | 21 (42.9) | 27 (51.9) | 27 (45.8) | 96 (45.1) |
| Total | 53 | 49 | 52 | 59 | 213 |
| Complete Pathological Response |  |  |  |  |  |
| No response | 62 (100.0) | 61 (96.8) | 63 (100) | 126 (98.4) | 249 (98.4) |
| Response | 0 (0) | 2 (3.2) | 0 (0) | 2 (1.6) | 4 (1.6) |
| Total | 62 | 63 | 63 | 128 | 253 |
| Type of Surgery |  |  |  |  |  |
| Breast conserving | 48 (73.8) | 47 (74.6) | 51 (77.3) | 51 (76.1) | 197 (75.5) |
| Mastectomy | 17 (26.2) | 16 (25.4) | 15 (22.7) | 16 (23.9) | 64 (24.5) |
| Total | 65 | 63 | 66 | 67 | 261 |
| Axillary Lymph Node Involvement at Surgery |  |  |  |  |  |
| No | 18 (45.0) | 26 (65.0) | 24 (57.1) | 23 (53.5) | 91 (55.2) |
| Yes | 22 (55.0) | 14 (35.0) | 18 (42.9) | 20 (46.5) | 74 (44.8) |
| Total | 40 | 40 | 42 | 43 | 165 |

**Table 4.** Rates across treatment groups.

| | Placebo | Celecoxib | $\chi^2$ | P |
| --- | --- | --- | --- | --- |
| **Objective Ultrasound-determined Response** | 41.2% (42/102) | 48.7% (54/111) | 0.916 | 0.339 |
| 95% CI (%) | 31.5, 51.4 | 39.1, 58.3 |  |  |
| **Complete Pathological Response** | 1.6% (2/125) | 1.6% (2/128) | <0.000 | 0.981 |
| 95% CI (%) | 0.2, 5.7 | 0.2, 5.5 |  |  |
| **Breast Conserving Surgery** | 74.2% (95/128) | 76.7% (102/133) | 0.103 | 0.749 |
| 95% CI (%) | 65.7, 81.5 | 68.6, 83.6 |  |  |
| **Axillary Lymph Node Involvement at Surgery** | 55.0% (44/80) | 55.3% (47/85) | 0.004 | 0.984 |
| 95% CI (%) | 43.5, 66.2 | 44.1, 66.1 |  |  |

Median follow-up was 5.1 years (range 0.1-7.1); loco-regional recurrence was reported in six patients in the AI+placebo group and seven patients treated with AI+celecoxib (Figure 2A). A total of 65 patients reported breast cancer progression in any location: 35 in the AI+placebo group, 30 in the AI+celecoxib group (Figure 2B). Progressive disease following initial breast cancer surgery was reported in 39 patients: 14 in the AI+placebo group, 25 in the AI+celecoxib group (χ^2^ = 0.549, *P* = 0.4). Overall survival rates were statistically similar in both groups: In total 42 deaths were reported; 17 in the AI+placebo group and 25 in the AI+celecoxib group (Figure 2C).

**Figure 2.**
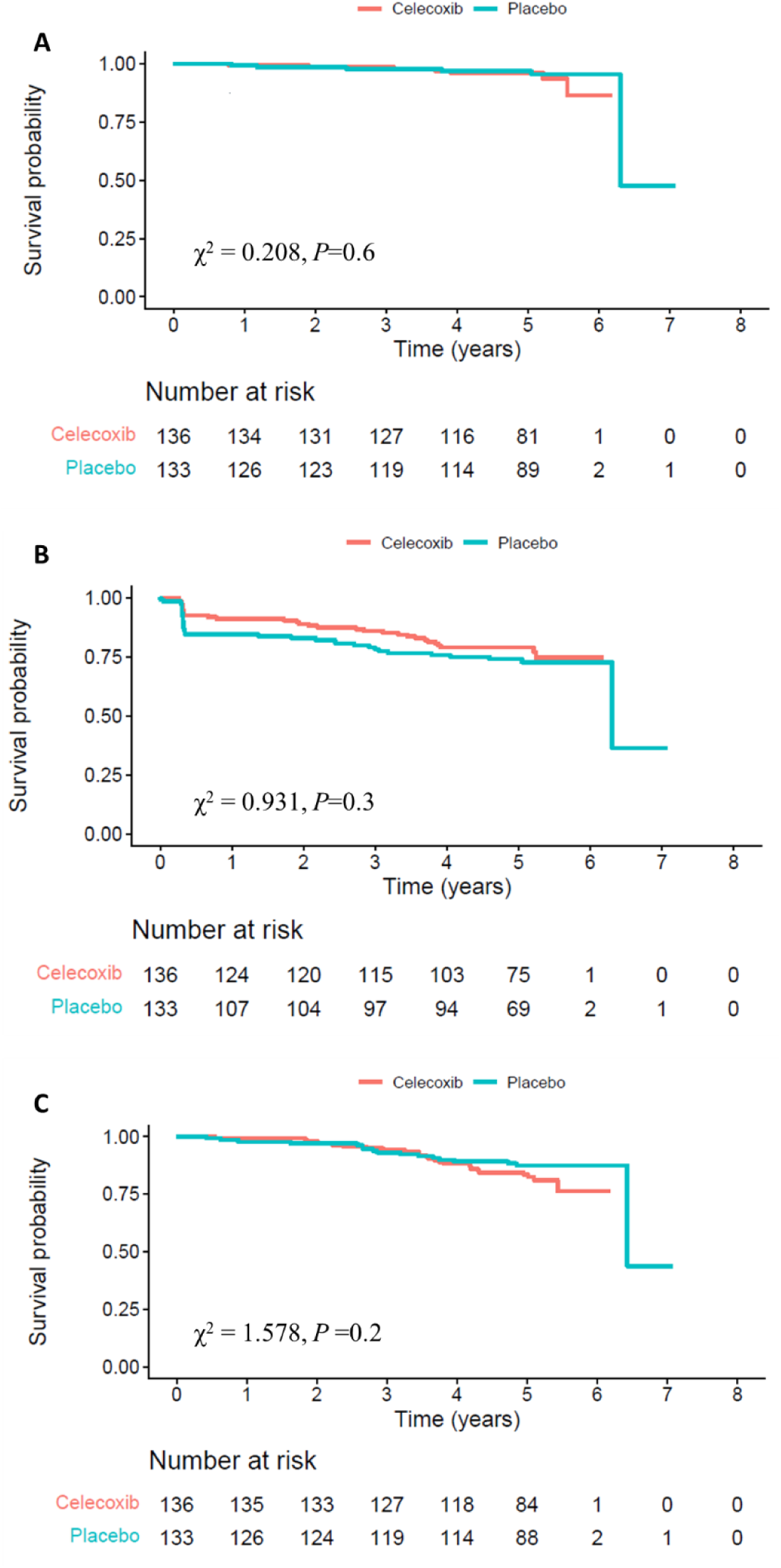
Time-to-event secondary outcomes. (A) Loco-regional recurrence free interval is shown, defined as time from trial entry date to date when local recurrence was first observed. Patients were censored at date last seen if an event was not reported. (B) Progression free recurrence interval is shown, defined as the time from the trial entry date to date when progression (tumour growth of greater than 20% between first and last assessment), relapse, or new primary tumour was observed. Patients were censored at date last seen if free of progression. (C) Overall survival is shown, defined as the time from trial entry date to date of deaths from any cause. Patients were censored at date last seen if still alive.

Seven-hundred and fifty-eight AEs were reported during NEO-EXCEL in 269 patients; 740 reported pre-surgery AEs and 18 reported post-surgery. The majority of AEs reported were grades 1-2 (703 [92.7%]) across all groups (Table 5). A list of all reported AEs according to treatment arm is provided in Appendix B.

**Table 5.** Total number of adverse events.

|  | Placebo |  | Celecoxib |  | Overall |
| --- | --- | --- | --- | --- | --- |
|  | Exemestane | Letrozole | Exemestane | Letrozole |  |
| Pre-surgery |  |  |  |  |  |
| Grade 1-2 | 192 | 145 | 185 | 167 | 689 |
| Grade 3-4 | 2 | 4 | 2 | 2 | 10 |
| Not Supplied | 1 | 3 | 3 | 2 | 9 |
| NA | 14 | 5 | 6 | 7 | 32 |
| Total pre-surgery | 209 | 157 | 196 | 178 | 740 |
| Post-surgery |  |  |  |  |  |
| Grade 1-2 | 7 | 1 | 6 | 0 | 14 |
| Grade 3-4 | 0 | 0 | 0 | 1 | 1 |
| NA | 3 | 0 | 0 | 0 | 3 |
| Total post-surgery | 10 | 1 | 6 | 1 | 18 |
| Total | 219 | 158 | 202 | 179 | 758 |

Note: No grade 5 adverse events were reported during the NEO-EXCEL trial.
NA, not assigned.

In total, 12 patients reported 13 serious adverse events during the trial, one cardiac arrhythmia was assessed as possibly related to trial treatment (Table 6).

**Table 6.** Categories of serious adverse events.

| Serious Adverse Event<br>Category | Placebo [*N* (%)] |  | Celecoxib [*N* (%)] |  | Overall [*N* (%)] |
| --- | --- | --- | --- | --- | --- |
|  | Exemestane | Letrozole | Exemestane | Letrozole |  |
| Cardiac Arrhythmia | 1 (33.3) | 1 (16.7) | 0 (0) | 0 (0) | 2 (13.3) |
| Haemorrhage/Bleeding | 0 (0) | 1 (16.7) | 2 (33.3) | 0 (0) | 3 (20) |
| Infection | 1 (33.3) | 2 (33.3) | 0 (0) | 0 (0) | 3 (20) |
| Musculoskeletal/Soft<br>Tissue | 1 (33.3) | 1 (16.7) | 0 (0) | 0 (0) | 2 (13.3) |
| NA | 0 (0) | 1 (16.7) | 0 (0) | 0 (0) | 1 (6.7) |
| Not supplied | 0 (0) | 0 (0) | 1 (16.7) | 0 (0) | 1 (6.7) |
| Pulmonary/Upper<br>Respiratory | 0 (0) | 0 (0) | 2 (33.3) | 0 (0) | 2 (13.3) |
| Renal/Genitourinary | 0 (0) | 0 (0) | 1 (16.7) | 0 (0) | 1 (6.7) |

NA, not assigned.

## Discussion

NEO-EXCEL is the first completed, phase III double-blind, placebo-controlled trial of the addition of celecoxib to AI as neoadjuvant endocrine therapy in early breast cancer. However, the primary outcome used (objective clinical response as measured by callipers) is a relatively crude method of assessment and this needs to be interpreted in the context of no benefit seen in any of the secondary outcomes reported. Due to slow recruitment during the initial stages of the trial a statistical redesign was introduced to reduce the required sample size. This trial demonstrated a significantly greater objective clinical response (CR+PR) when celecoxib was combined with AI, however, its results must still be interpreted with some caution. Particularly as this difference while trending in the same direction was smaller and not significant when measuring response radiologically using ultrasound. Long term outcome was unaffected with PFS and OS statistically similar between both groups. This suggests any benefit seen is limited to the neoadjuvant phase only; the placebo-controlled nature of this trial makes the likelihood of investigator bias unlikely. The clinical size change did not reduce the mastectomy rate. An anti-inflammatory effect of AI+celecoxib on peri-tumour tissue resulting in a clinical reduction in size when measured by calliper may explain these discordant findings. However, this is a speculative explanation as inter-observer heterogeneity of calliper measurements was not ascertained during study. Many patients in NEO-EXCEL were suitable for breast conservation at study entry. We cannot know if this study was confined to only those requiring downstaging to permit breast conservation as it would have impacted on surgical outcomes.

Within NEO-EXCEL pathological tumour size was measured post-operatively but no clinically relevant difference was observed (28.0mm Placebo/Exemestane; 26.0mm Placebo/Letrozole; 25.0mm Celecoxib/Exemestane; and 27.0mm Celecoxib/Letrozole). Furthermore, as no pre-operative size can be measured, changes in size via this method cannot be reported. Residual cancer burden score was not evaluated as an endpoint during the trial which is only validated in the context of neoadjuvant chemotherapy.

NEO-EXCEL also reports that addition of celecoxib to exemestane results in a greater reduction in tumour size than combining celecoxib and letrozole. This effect was unexpected and, although statistically significant, again should be interpreted with caution, particularly as exemestane and letrozole have previously been shown to have equivalent activity [4]. It may represent a genuine difference in the interactions of steroidal- and nonsteroidal-AIs with celecoxib, an anti-inflammatory agent, and may warrant further investigation. However, in this trial the numerical response rate to exemestane alone (49%) is lower than the response to letrozole (62%). Therefore, this could represent a chance numeric effect or may reflect uncaptured differences in initial tumour characteristics and/or an imbalance between the arms or an effect of a small subgroup e.g., in the exemestane+placebo group >10% more axillary node involvement at surgery was observed (Table 3).

The increased clinical response rate observed does not result in an improved rate of breast conservation. As only 33% of patients were considered unsuitable for breast conservation at trial entry, the trial is underpowered to demonstrate an improvement in breast conservation. In practice, down-staging tumour size is only one of several criteria, such as patient choice, tumour position, and presence of extensive malignant microcalcification, used to decide the scope of surgery required after neoadjuvant therapy.

We note that within NEO-EXCEL, there was a high proportion of patients with ER+HER2+ tumours (34.9%). This most likely reflects case selection with larger tumours more likely to be HER-2 positive. Neoadjuvant anti HER-2 based therapy has now become standard of care in this group and was not available or in use during recruitment to this study.

There have been a small number of studies evaluating the addition of celecoxib to AIs in women with operable HR+ breast cancer. In the Celecoxib Anti-Aromatase Neoadjuvant (CAAN) trial Chow and colleagues included 82 postmenopausal women with a positive ER and/or progesterone receptor status randomised three ways to exemestane 25mg od in combination with celecoxib 400mg bid, or exemestane 25mg od, or letrozole 2.5mg od. The objective clinical response rate measured by callipers was 58.6% for the combination, 54.5% for exemestane, and 62.0% for letrozole; differences between groups were not significant. A smaller study investigating biomarker changes in postmenopausal women with early HR+ breast cancer included 22 women treated with exemestane 25mg od for eight weeks, followed by exemestane 25mg od combined with celecoxib 400mg bid for eight weeks. ER, PR, Ki-67, and COX-2 protein expression decreased significantly but, contrary to expectation, no significant changes in aromatase expression were found with the addition of celecoxib to exemestane in this study [23]. Unfortunately, sample quality issues prevented any tumour gene expression analyses in this study.

Within the adjuvant setting, the REACT study evaluated the role of celecoxib at a lower dose of 400mg od for patients with ER+ breast cancer. This study concluded that two years of celecoxib treatment did not improve DFS [24]. The authors consider whether a higher dose or longer duration would lead to a DFS benefit. NEO-EXCEL does not lend support to the exploration of a higher dose; however, prolonged duration remains an unanswered question. Indeed, as 400mg bid AI-celecoxib combination therapy is well-tolerated, treatment for longer than 16 weeks may be of greater clinical benefit.

Clinical response remains an important endpoint in comparing the effectiveness of the AI+celecoxib combination in this setting with other contemporaneous studies. There is increasing interest in utilising Ki67 levels as a surrogate marker of neo-adjuvant success in breast cancer. Whilst it has been shown that Ki67 decreases with AI use, this should be assessed with particular caution when assessing the impact of celecoxib. The mechanism of action of celecoxib is not likely mediated through Ki67 so it is inappropriate to use the preoperative endocrine prognostic index (PEPI) score in this context. Again, no significant change in Ki67 was detected by Martin et al when examining 23 paired sets of biopsies in a celecoxib versus no treatment population. The Ki67 change in the cohort given celecoxib was - 16% (95% CI −31.4-+1.4; *P*=0.056), and in the no treatment group was −8.1 (95% CI- −23.2-+10.1; *P*=0.24). There is no statistical difference between the two groups (*P*=0.45).

The greatest strength of NEO-EXCEL is its prospective, double-blind, placebo-controlled design. However, it also carries limitations that reduce the impact of its results. The primary assessment method of clinical response may be impacted by peritumoral tissue effects and may explain the positive primary endpoint result with negative secondary endpoints. Slow recruitment of the trial within the first two years of opening also hampered the trial’s goal of being a definitive phase III trial and ultimately precluded the originally planned analyses to detect any differences between exemestane and letrozole. Recruitment was delayed by many factors including site ethical and R&D approval, and the set-up of pharmacies. At the time this was common amongst most of the breast cancer trials in the UK but exacerbated by the original ambition of this trial to be the largest neoadjuvant trial in the UK requiring 70 sites to open. Recruitment during NEO-EXCEL was also limited by the lack of required trial experience and research training of UK surgeons many were unable to act as local lead investigators, a challenge to the set-up of this trial, which was not anticipated and a situation which has now been largely resolved partly as a result of the NEO-EXCEL experience. Finally, the delay in reporting of the trial has also negatively impacted the relevance of the results given the changes in neoadjuvant and adjuvant therapy that have emerged since the start of this study, and we also acknowledge that there was no patient or public involvement in the design, conduct or reporting of the trial due to the time period in which it was conducted.

While there are interesting aspects to explore such as the potential increased effect of celecoxib with exemestane, the future of clinical COX-2 endocrine therapy research is currently on hold as there are many alternative approaches, which hold out greater promise. The spectre of cardiac toxicity from long term COX-2 treatment is ever present, but it is reassuring that this study reports no significant cardiac toxicity from short exposure to higher dose celecoxib and the REACT study has demonstrated the cardiac safety of a long-term lower dose celecoxib in early breast cancer patients.

### Conclusions

Although hampered by slow recruitment reducing the initial ambition of the trial and the NEO-EXCEL remains the first completed, phase III double-blind, placebo-controlled trial of the addition of celecoxib to AI as neoadjuvant endocrine therapy in early breast cancer. Clinical response improvement with a lack of significant ultrasound-determined response improvement or any surgical or long-term outcomes benefit, however, did not provide clearcut evidence of AI+COX-2 inhibition improving treatment for ER+ early resectable postmenopausal breast cancers.

## Data Availability

All data produced in the present study are available upon reasonable request to the authors

## List of Abbreviations

AE: adverse event
AI: aromatase inhibitor
bid: twice daily
CI: confidence intervals
COX-2: cyclooxygenase-2
CR: complete response
CTCAE: Common Terminology Criteria for Adverse Events
DFS: disease-free survival
DMC: data monitoring committee
ER+: oestrogen-receptor positive
HER: human epithelial growth factor receptor
HR+: hormone receptor positive
IQR: interquartile range
NCI: National Cancer Institute
od: once daily
OR: odds ratio
OS: overall survival
PFS: progression-free survival
PR: partial response
RECIST: Response Evaluation Criteria In Solid Tumours

## Data availability

Participant data and the associated supporting documentation will be available within 6 months after the publication of this manuscript. Details of our data request process is available on the CRCTU website. Only scientifically sound proposals from appropriately qualified research groups will be considered for data sharing. The decision to release data will be made by the CRCTU Director’s Committee, who will consider the scientific validity of the request, the qualifications and resources of the research group, the views of the Chief Investigator and the trial steering committee, consent arrangements, the practicality of anonymising the requested data and contractual obligations. A data sharing agreement will cover the terms and conditions of the release of trial data and will include publication requirements, authorship and acknowledgements and obligations for the responsible use of data. An anonymised encrypted dataset will be transferred directly using a secure method and in accordance with the University of Birmingham’s IT guidance on encryption of data sets.

## Conflicts of interest

RCS was supported by the National Institute for Health Research University College London Hospitals Biomedical Research Centre. CJP received an honorarium from Pfizer for a lecture given and participated in an Advisory Board for Pfizer. AF, DWR, JMSB, PAC, and JAD received research funding from Pfizer and Cancer Research UK to fund this study. All remaining authors have declared no conflicts of interest.

## Funding statement

This work was supported by Cancer Research UK (C11924/A5683); CRUK trial number: CRUK/06/005. Pfizer provided an educational grant for the trial and supplied the study medication, celecoxib, free of charge.

Staff at the CRCTU are supported by a core funding grant from Cancer Research UK (CTUQQR-Dec22/100006).

The trial was initiated and conducted independently by the trial investigators. The funders had no role in trial design, data collection, data analysis, data interpretation or writing of the report.

The corresponding author had full access to all the data in the trial and had final responsibility for the decision to submit for publication.

## Authors’ contributions

AF and DWR designed the trial, interpreted data, and wrote the manuscript; CLB and CJP designed the trial; JAD designed the trial and interpreted data; AP and SP produced the analysis and figures; KH wrote the manuscript; JMSB, MG, ES, CG, and SJB provided sponsor and trial management oversight; AF, CP, RCS, PAC, CJP, ARP, and DWR recruited patients to the study. All authors critically reviewed the manuscript.

## Acknowledgements

We acknowledge the 42 investigators and their teams from 22 participating UK centres who entered patients into the NEO-EXCEL trial; a full list can be found in Appendix C. Our gratitude also goes to the 269 women who kindly participated in our study. We thank the Data Monitoring Committee members: Jon Olson, Bent Ejlertsen (Rishopitalet, Copenhagen), Roger A’Hern (The Institute of Cancer Research, London), Laura Buckley (University of Birmingham), Michael Douek (Guy’s Hospital, London), Josephine Khan (University of Birmingham), Shalini Chaudhri, Isabelle Hero and Jane Steven (Queen Elizabeth Hospital, Birmingham), John Robertson (Nottingham City Hospital) and Mike Hallissey (Queen Elizabeth Hospital, Birmingham). We thank the trials staff at the coordinating centre - Cancer Research UK Clinical Trials Unit (CRCTU), University of Birmingham, UK; and the translational centre – Biomarkers and Companion Diagnostics, University of Edinburgh, UK; and Maren White, Appletree Medical Writing, for support in the production of this manuscript; and Dr Siân Lax (CRCTU, University of Birmingham) for contributions to the paper.

Preliminary results from NEO-EXCEL were presented in part at the San Antonio Breast Cancer Symposium, December 2015 [25].

## Appendix A

**Table A.1:**
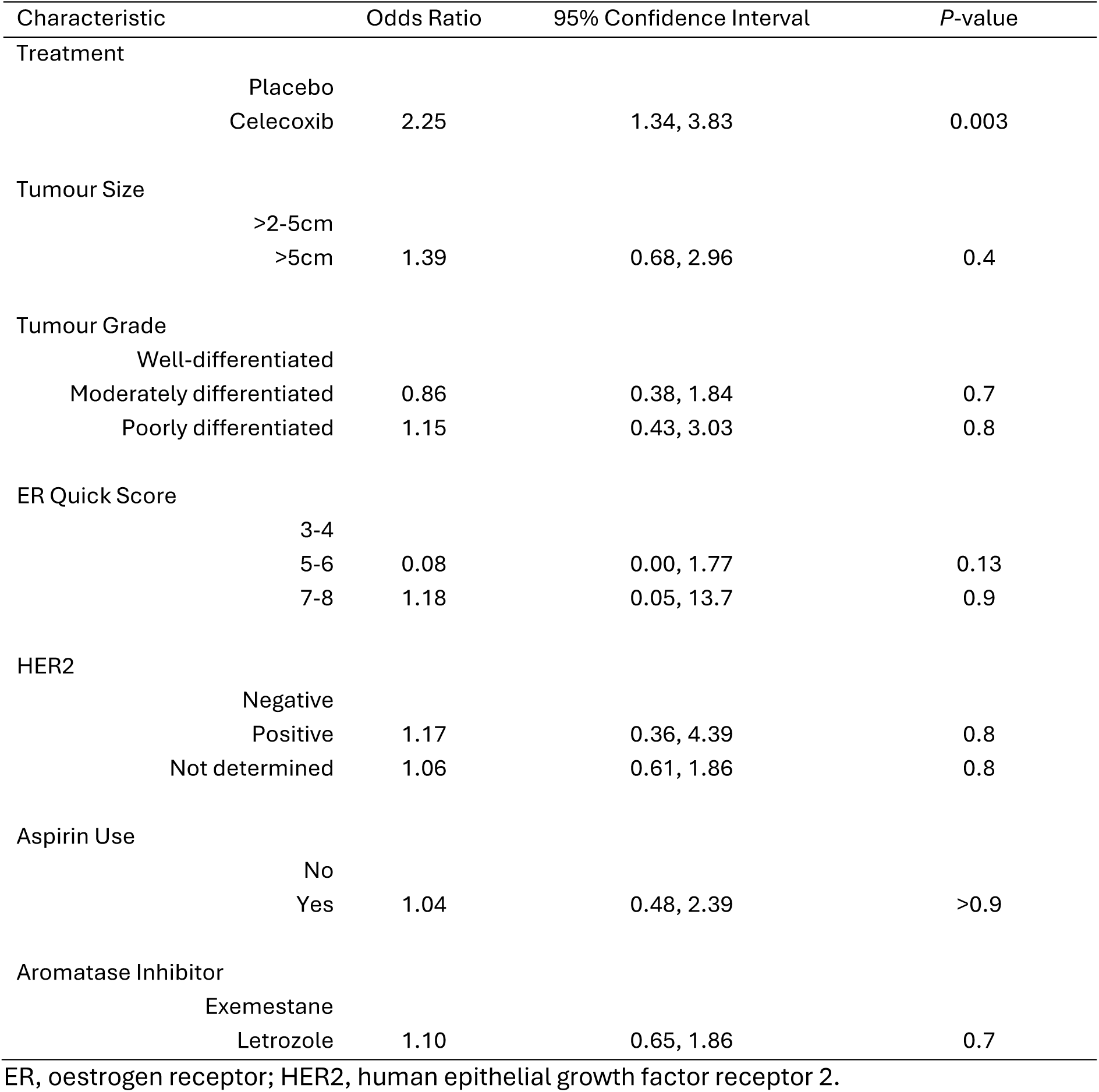
Multivariate logistic regression model.

| Characteristic |  | Odds Ratio | 95% Confidence Interval | P-value |
| --- | --- | --- | --- | --- |
| Treatment |  |  |  |  |
|  | Placebo |  |  |  |
|  | Celecoxib | 2.25 | 1.34, 3.83 | 0.003 |
| Tumour Size |  |  |  |  |
|  | >2-5cm |  |  |  |
|  | >5cm | 1.39 | 0.68, 2.96 | 0.4 |
| Tumour Grade |  |  |  |  |
|  | Well-differentiated |  |  |  |
|  | Moderately differentiated | 0.86 | 0.38, 1.84 | 0.7 |
|  | Poorly differentiated | 1.15 | 0.43, 3.03 | 0.8 |
| ER Quick Score |  |  |  |  |
|  | 3-4 |  |  |  |
|  | 5-6 | 0.08 | 0.00, 1.77 | 0.13 |
|  | 7-8 | 1.18 | 0.05, 13.7 | 0.9 |
| HER2 |  |  |  |  |
|  | Negative |  |  |  |
|  | Positive | 1.17 | 0.36, 4.39 | 0.8 |
|  | Not determined | 1.06 | 0.61, 1.86 | 0.8 |
| Aspirin Use |  |  |  |  |
|  | No |  |  |  |
|  | Yes | 1.04 | 0.48, 2.39 | >0.9 |
| Aromatase Inhibitor |  |  |  |  |
|  | Exemestane |  |  |  |
|  | Letrozole | 1.10 | 0.65, 1.86 | 0.7 |

ER, oestrogen receptor; HER2, human epithelial growth factor receptor 2.

## Appendix B

**Table B.1:**
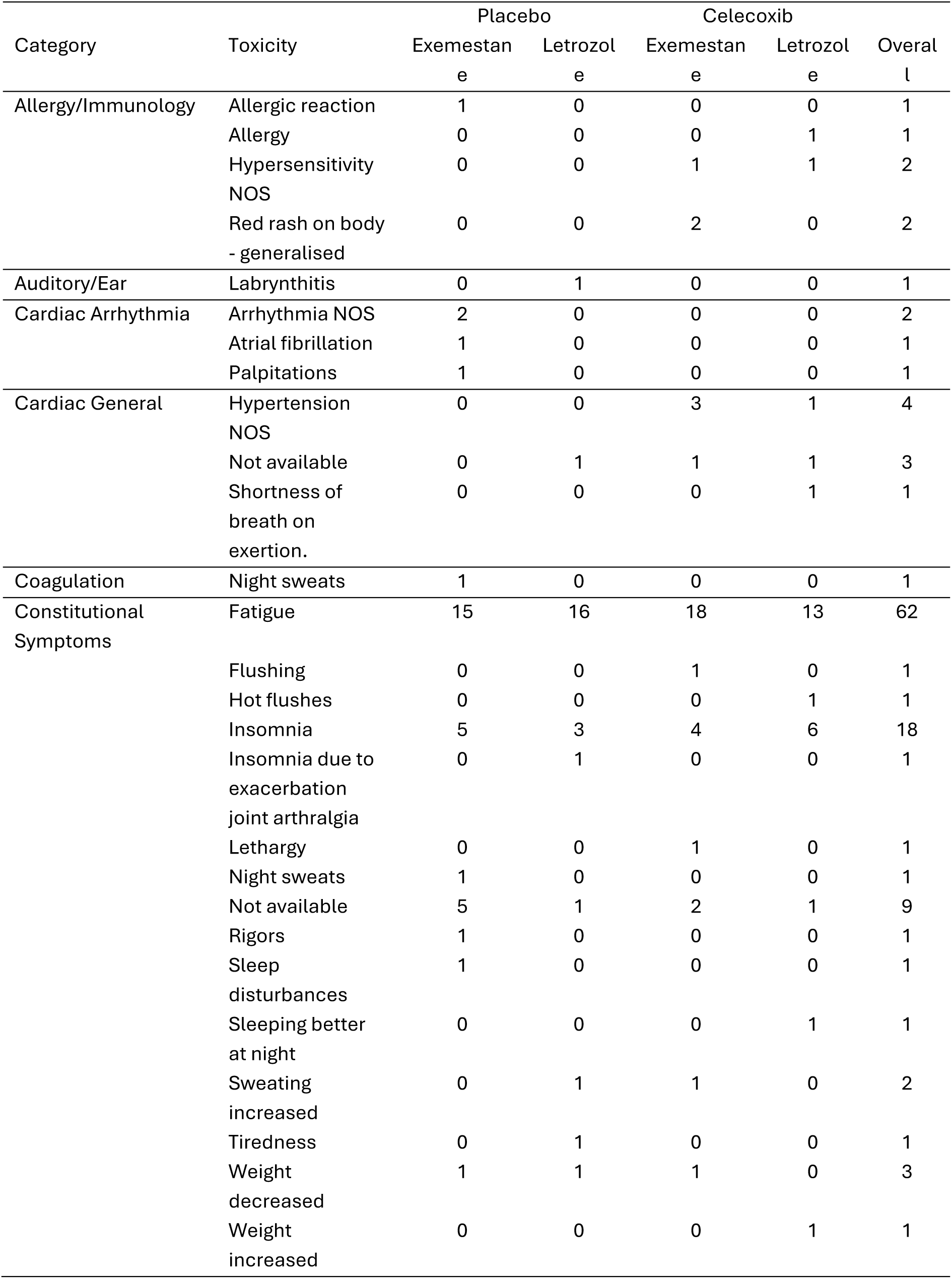
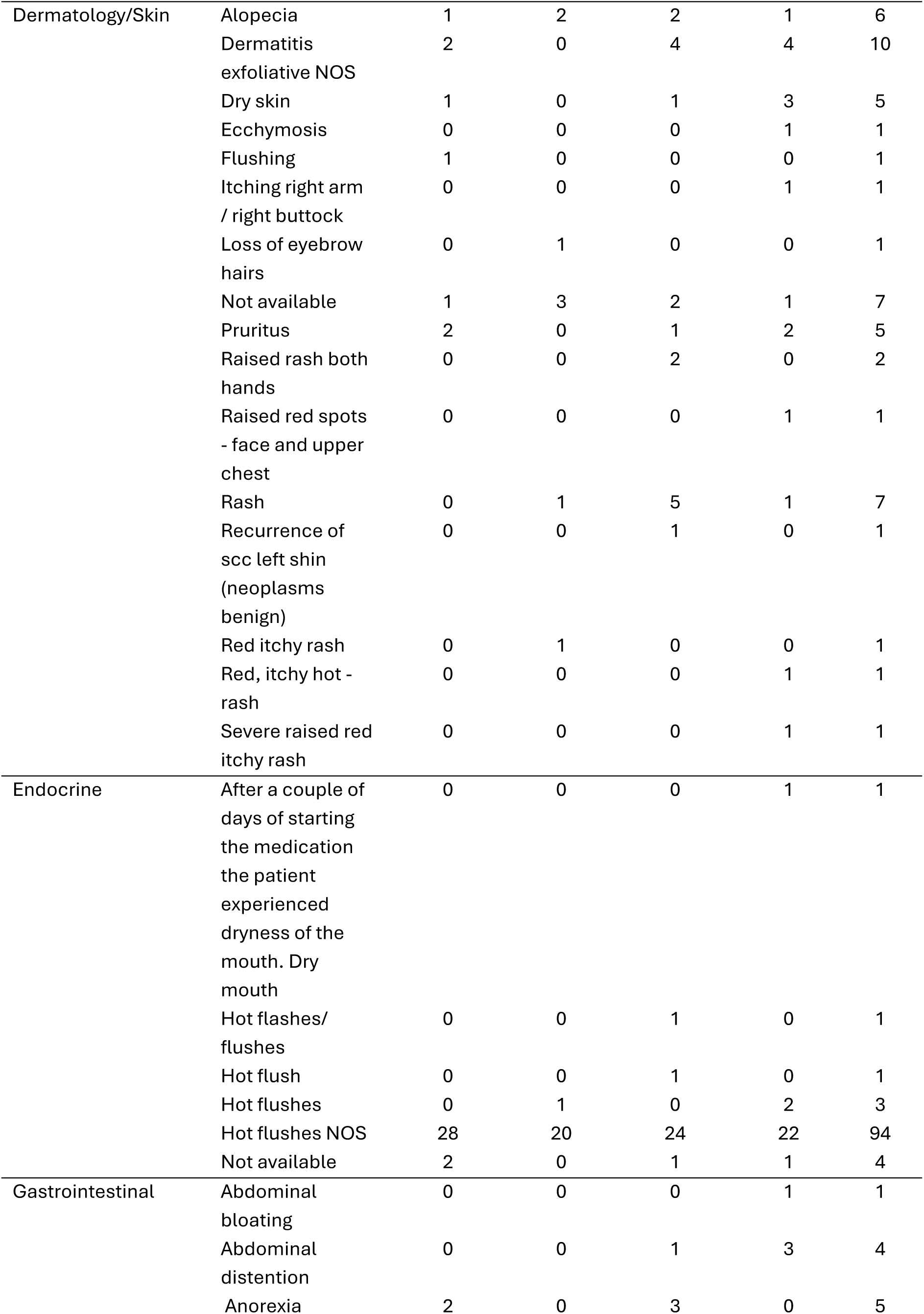
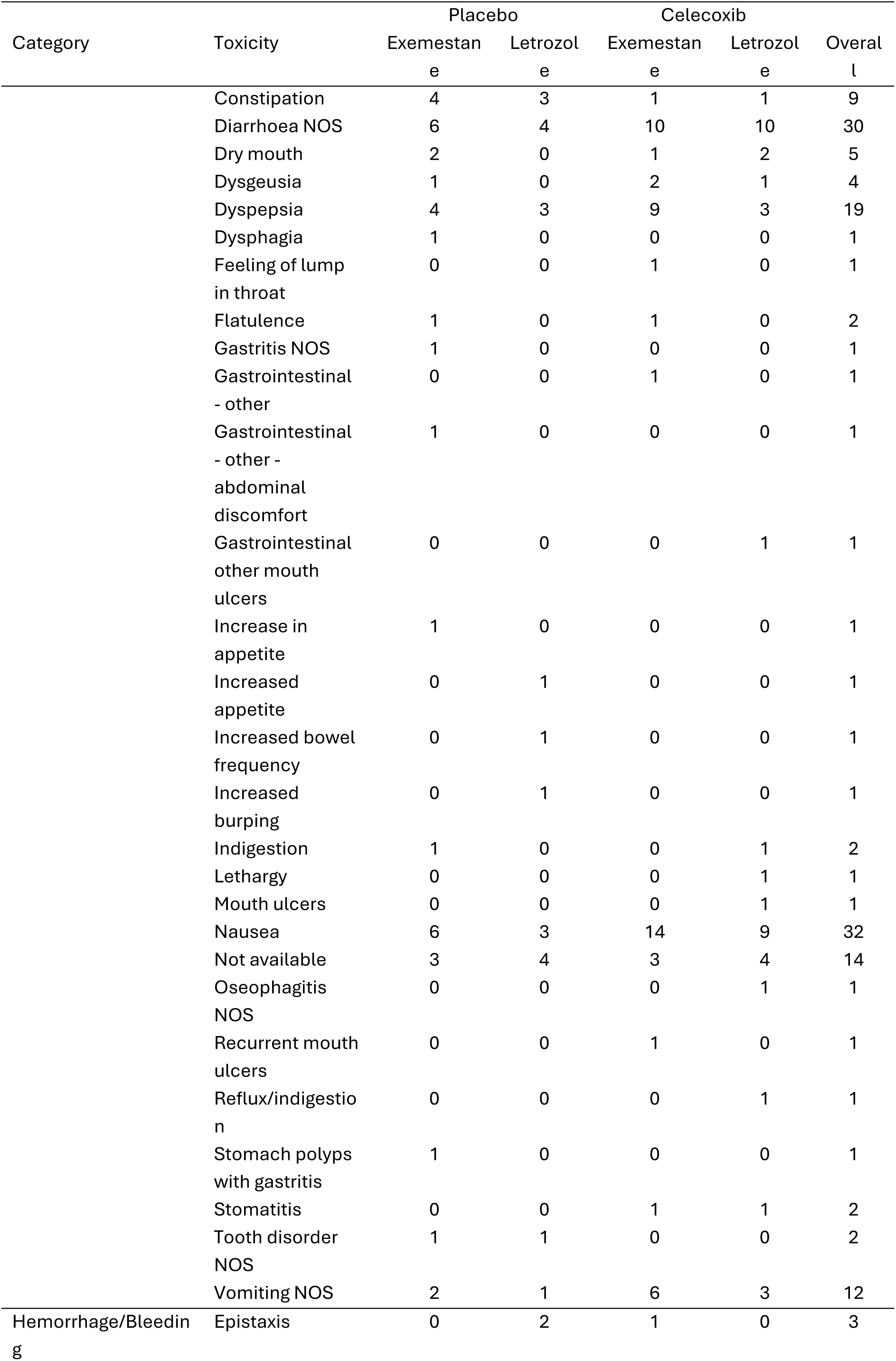
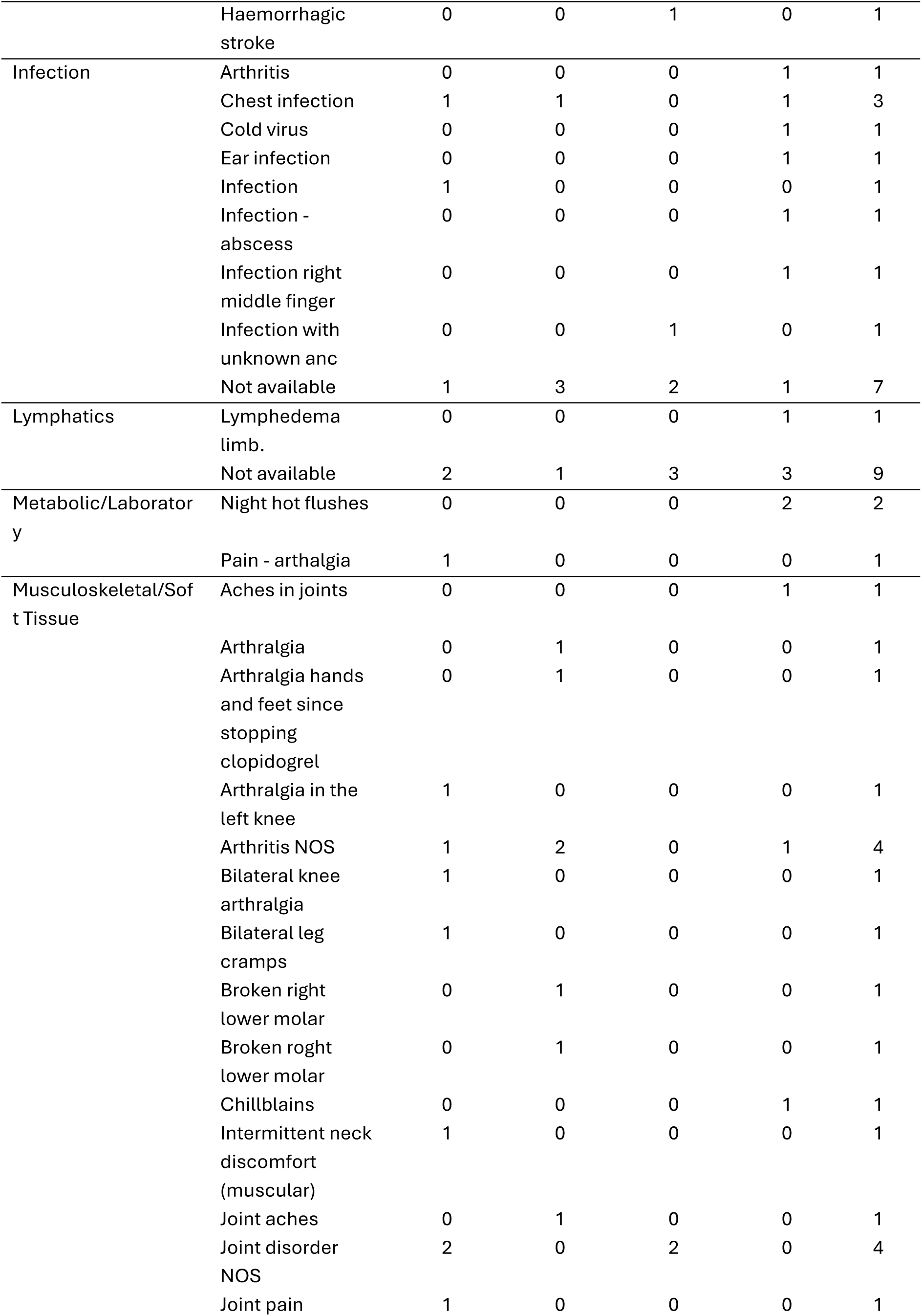
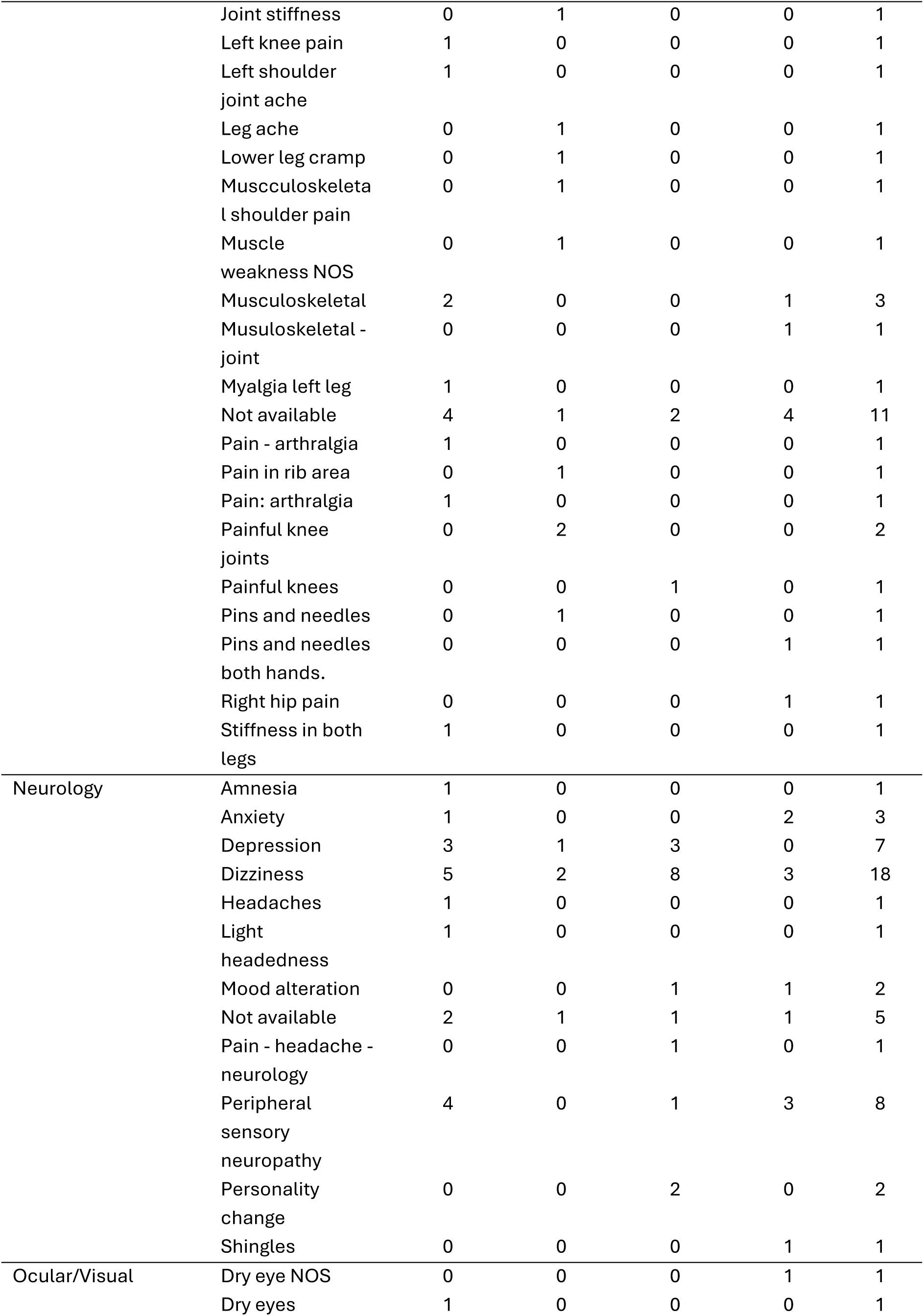
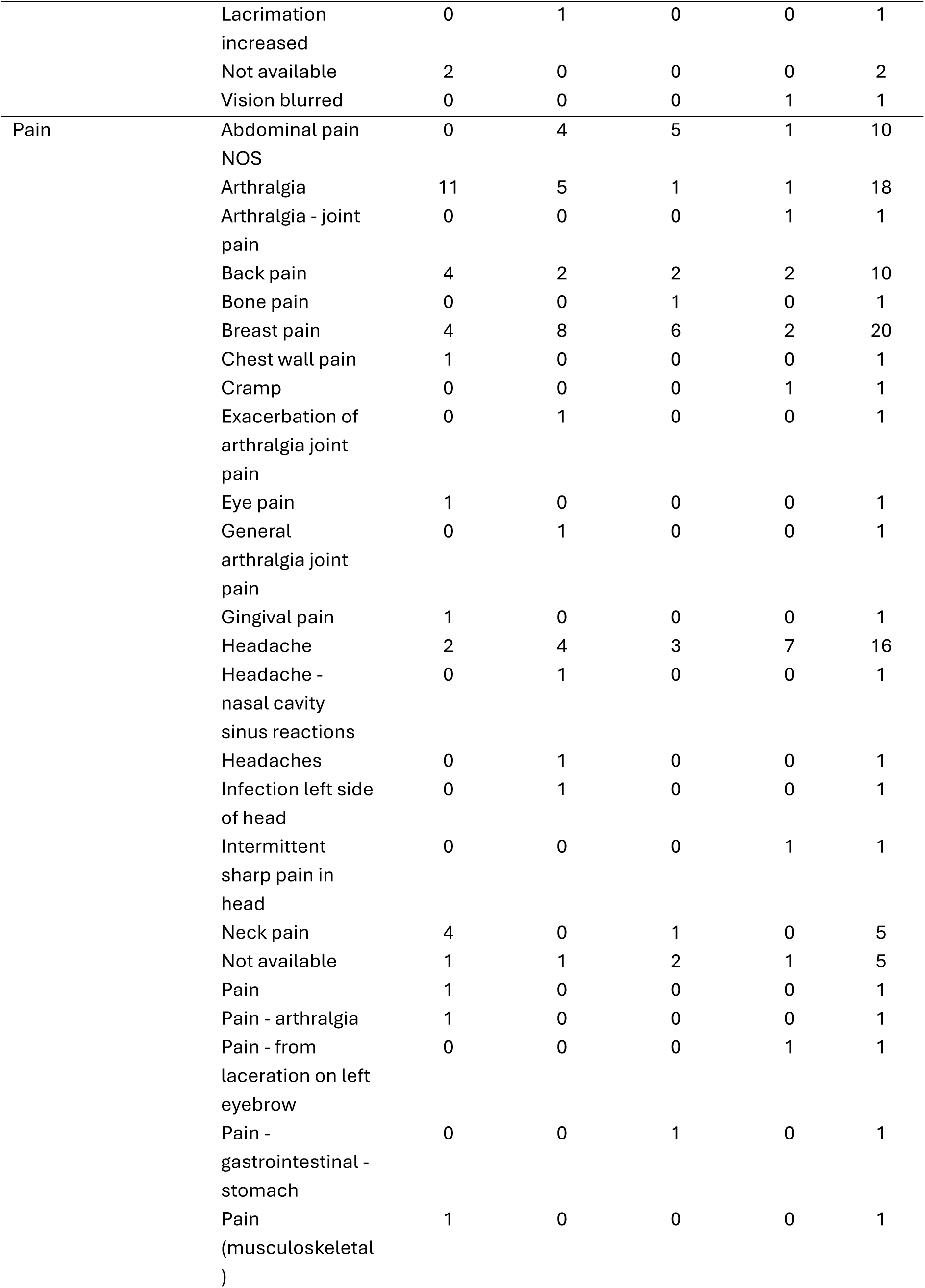
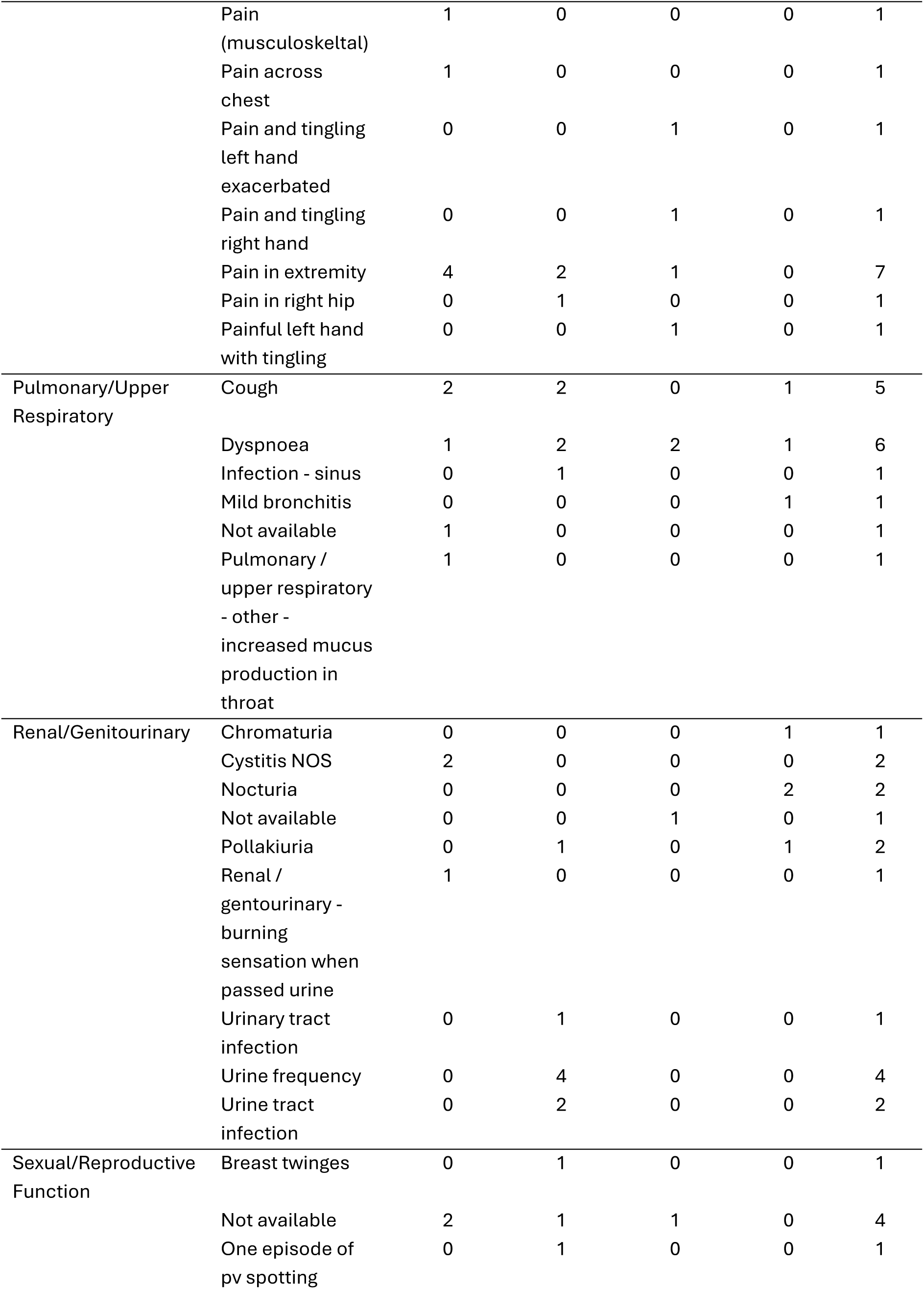
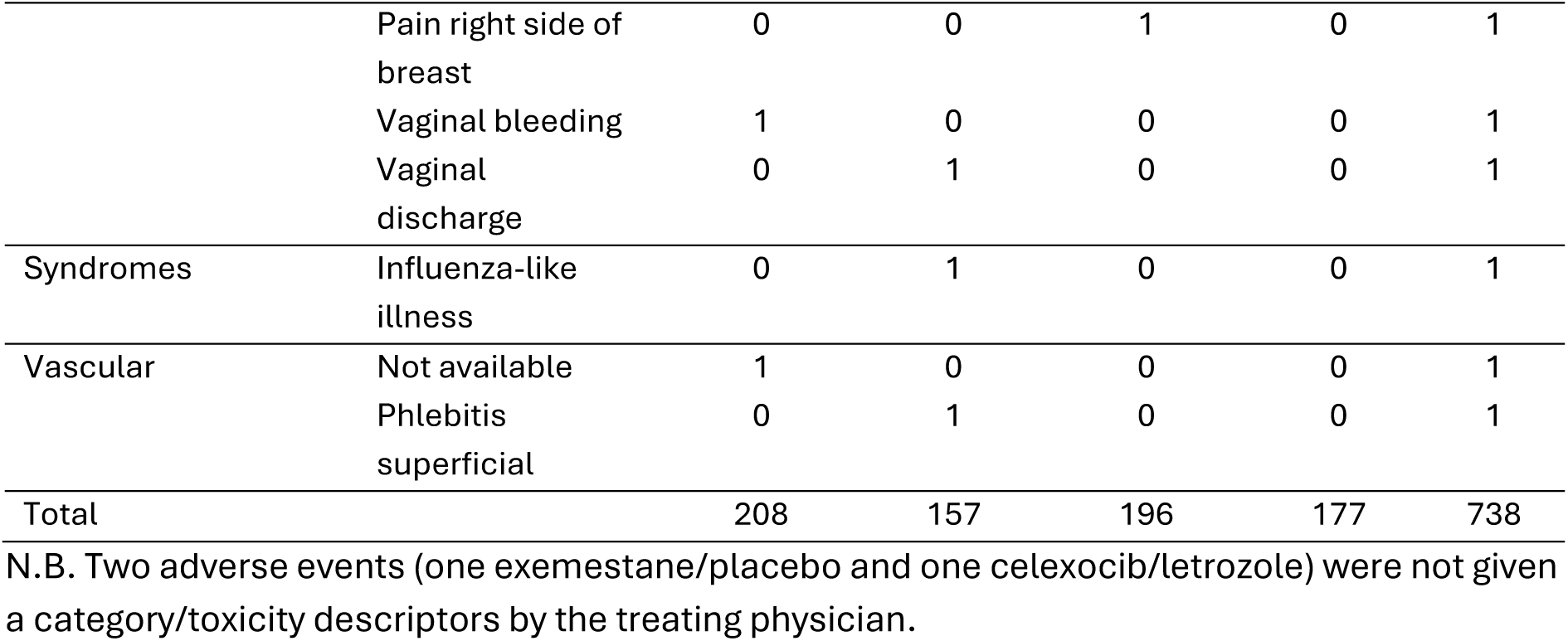
Number of pre-surgery adverse events split by treatment group.

**Table B.2:** Number of post-surgery adverse events split by treatment group.

| Category | Toxicity | Placebo |  | Celexocib |  | Overall |
| --- | --- | --- | --- | --- | --- | --- |
|  |  | Exemestane | Letrozole | Exemestane | Letrozole |  |
| Constitutional Symptoms | Fatigue | 0 | 0 | 1 | 0 | 1 |
|  | Insomnia | 1 | 0 | 0 | 0 | 1 |
| Gastrointestinal | Dry mouth | 0 | 0 | 1 | 0 | 1 |
|  | Nausea | 1 | 0 | 0 | 0 | 1 |
| Hemorrhage/Bleeding | Haematoma | 1 | 0 | 0 | 0 | 1 |
|  | Post procedural haemorrhage | 1 | 0 | 0 | 0 | 1 |
| Infection | Not available | 1 | 0 | 0 | 0 | 1 |
|  | Wound discharge | 0 | 0 | 0 | 1 | 1 |
|  | Wound infection | 0 | 0 | 1 | 0 | 1 |
| Lymphatics | Not available | 1 | 0 | 0 | 0 | 1 |
|  | Oedema Limb | 1 | 0 | 0 | 0 | 1 |
| Musculoskeletal/Soft Tissue | Fall / Back Pain | 0 | 0 | 1 | 0 | 1 |
|  | Seroma | 0 | 0 | 1 | 0 | 1 |
| Neurology | Anxiety | 1 | 0 | 0 | 0 | 1 |
| Pulmonary/Upper Respiratory | Cough | 0 | 1 | 0 | 0 | 1 |
| Sexual/Reproductive Function | Pain in left side of breast | 0 | 0 | 1 | 0 | 1 |
| Total |  | 8 | 1 | 6 | 1 | 16 |

N.B. Two adverse events (both exemestane/placebo) were not given a category/toxicity descriptors by the treating physician.

## Appendix C

A full list of the participating UK centres who entered patients into the NEO-EXCEL trial:

- Frenchay Hospital, Dr Mike Shere
- Southport and Formby District General Hospital, Mr Sabah Jmor
- St James’s University Hospital, Mr Raj Achuthan
- Grantham and District Hospital, Mr Jibril Jibril
- City Hospital, Prof Daniel Rea
- Good Hope Hospital, Mr Tahir Farooq
- The Queen Elizabeth Hospital, Prof Adele Francis
- Royal United Hospital, Mr Richard Sutton
- Cheltenham General Hospital, Mr James Bristol
- Barnet Hospital, Mr Muhamed Al Dubaisi
- Frimley Park Hospital, Mr Raouf Doaud
- Broomfield Hospital, Mr Simon Smith
- Chelmsford and Essex Centre, Prof Paul Sauven
- Essex County Hospital, Mr Sankaran Chandrasekharan
- Princess Royal University Hospital, Mr Pankesh Sinha
- Leeds General Infirmary, Mr Mark Lansdown
- Wythenshawe Hospital, Prof Nigel Bundred
- St Margaret’s Hospital, Mr Ashraf Patel
- Wishaw General Hospital, Mrs Alison Lannigan
- University Hospital, Coventry, Prof Christopher Poole
- Forth Valley Royal Hospital, Dr Judith Fraser
- Peterborough City Hospital, Mr Steven Goh

## Notes

### Competing Interest Statement

The authors have declared no competing interest.

### Clinical Trial

EudraCT Number: 2006-000436-27
ISRCTN number: 09768535

### Clinical Protocols

https://www.birmingham.ac.uk/documents/college-mds/trials/crctu/neo-excel/forinvestigators/neoexcel-protocol-9.0-vd09-nov-2018.pdf

### Author Declarations

West Midlands Medical Research Ethics Committee, Birmingham, England, gave ethical approval for this work.

